# A systematic literature review of methodological approaches, challenges, and opportunities in the application of Mendelian randomisation to lifecourse epidemiology

**DOI:** 10.1101/2023.05.16.22283780

**Authors:** Grace M. Power, Eleanor Sanderson, Panagiota Pagoni, Abigail Fraser, Tim Morris, Claire Prince, Timothy M. Frayling, Jon Heron, Tom G. Richardson, Rebecca Richmond, Jessica Tyrrell, George Davey Smith, Laura D. Howe, Kate Tilling

## Abstract

**Background:** Diseases diagnosed in adulthood may have antecedents throughout – including prenatal – life. Gaining a better understanding of how exposures at different stages in the lifecourse influence health outcomes is key to elucidating the potential benefits of specific disease prevention strategies. However, confounding is highly likely in studies with earlier life or time-varying exposures. Mendelian randomisation (MR) is therefore increasingly used to estimate causal effects of exposures across the lifecourse on later life outcomes.

**Methods:** This systematic literature review aims to identify MR methods used to perform lifecourse investigations and review previous work that has utilised MR to elucidate the effects of factors acting at different stages of the lifecourse. We conducted a systematic search in PubMed, Embase, Medline and MedRXiv databases to comprehensively obtain lifecourse epidemiology studies that have employed MR.

**Results:** Thirteen methodological studies were identified. Four studies focused on the impact of time-varying exposures on the interpretation of “standard” MR techniques, five presented methods for analysing repeat measures of the same exposure, and four described novel methodological approaches to handling parental exposures in relation to offspring outcomes. A further 84 studies presented the results of an applied research question with relevance to lifecourse epidemiology. Over half of these estimated effects in a single generation and were largely confined to the exploration of questions regarding body composition. Of the one generational studies employed in this review, 59% estimated the effect of exposures at birth, birth to/and childhood, birth to/and adolescence or birth to/and adulthood, 30% at childhood, childhood to/and adolescence or childhood to/and adulthood, and 11% at adolescence or adulthood. The remaining looked across two generations. These estimated effects of maternal exposures, with one study additionally examining paternal exposures, in relation to offspring outcomes.

**Conclusion:** There is a growing body of research focused on the development and application of MR methods to address lifecourse research questions. The possibility that genetic effects have different levels of importance in the progression of an exposure at different ages should be more commonly considered for application in an MR context. Limitations exist, however, specifically regarding data constraints.

## Introduction

Diseases diagnosed in adulthood often have antecedents throughout – including prenatal – life (1). A lifecourse approach recognises the contribution of physical or social exposures acting during gestation, childhood, adolescence, and earlier or adult life or across generations to long-term biological, behavioural, and psychosocial processes that link adult health and disease risk (2, 3). Gaining a better understanding of how exposures at different stages in the lifecourse influence health outcomes is key to elucidating the potential benefits of specific disease prevention strategies. However, confounding is highly likely in studies with earlier life and time-varying exposures and later life health outcomes (4), particularly time-varying confounding and intermediate (or mediator-outcome) confounding (5). Intergenerational and family level factors may also contribute to further distinctive sources of confounding in multigenerational studies. Approaches to interrogate causality by minimising confounding are therefore of importance to strengthen causal inference in a lifecourse setting (6, 7).

Mendelian randomisation (MR) exploits the random assortment of genetic variants, independent of other traits, to enable analyses that largely mitigate against distortions resulting from confounding and reverse causality (8). This is a key motivation behind using a MR approach, which estimates the causal effect of modifiable risk factors under three assumptions; the instrumental variables used must i) be associated with the exposure of interest (‘relevance’), ii) not share common causes with the outcome (‘independence’ or ‘exchangeability’) and iii) not affect the outcome other than through the exposure (‘exclusion’). Several statistical methods have been proposed for MR with individual-level as well as summarised data. In a one-sample setting with individual-level data, a causal effect estimate is often obtained using the two-stage least-squares (2SLS) method (9). It is more common for two-sample investigations to use summarized data. In addition, at the introduction of MR, it was recognised that the association of genetic variants with exposures could change with age, which needed to be taken into account in interpretation (10, 11).

The application of MR to lifecourse research questions has two key challenges. Firstly, we are interested in isolating the causal effects of age-specific exposures. MR studies typically use a single measurement of an exposure to estimate its effects on an outcome (henceforth termed “standard” MR) and genes are invariable across the lifecourse. As such, results obtained are often interpreted as the average lifetime effect of the genetically predicted exposure, or genetic liability for an exposure if that exposure is binary (12). Whilst this approach is sufficient for some exposures, it requires extension to address lifecourse questions. This extension is possible in cases where inherited genetic variants have different effects at different timepoints in the lifecourse (within a population), allowing us to separate time-varying effects of certain exposures (13–15). Secondly, some lifecourse research questions involve the exploration of parental exposures. The inclusion of multiple generations brings additional analytical and methodological challenges due to common confounding and genetic relatedness.

This systematic literature review has two core aims. Firstly, to identify MR methods that have been developed to evaluate or conduct lifecourse epidemiological investigation and secondly, to systematically review previous work that has utilised MR to elucidate the impacts of risk factors from different stages of the lifecourse on later life outcomes. These studies fulfil the criteria outlined in the STROBE-MR guidelines, and specifically respond to the criterion of whether effect estimates previously derived would generalise to other exposure periods (16, 17).

## Methods

### 1. Search strategy and eligibility criteria

The protocol for this systematic literature review was registered in the International Prospective Register of Systematic Reviews (PROSPERO) as CRD42022314287 and was conducted in line with the 2020 Preferred Reporting Items for Systematic Reviews and Meta-Analyses (PRISMA) guidelines (18). We searched for lifecourse epidemiology studies, defined as studies investigating biological, behavioural, and psychosocial processes that link health and disease risk to exposures that take place in a preceding life stage. For example, studies relating pre-gestation, gestation, early life, childhood, or adolescence exposures to adult outcomes would be included. Studies linking adult exposures to adult outcomes would be included if the adult exposure is related to a particular stage/phase of adulthood, such as menopause, as long as this stage/phase predates the outcome under study. We also include in our definition of lifecourse studies the effects of repeated measures of the same time-varying exposure on a later outcome (See Supplementary Material 1) (19). Studies were eligible from any geographical location, with individuals from any age group and which included a MR study design (i.e., a study using genetic variants to determine whether there is a causal relationship between a modifiable risk factor and an outcome). We include as an “MR study” any study that uses genetic variants related to an exposure of interest to understand the causal nature of the relationship between that exposure and an outcome of interest. This includes studies where the genetic variants are used as an instrumental variable, and those where the association between the genetic variants and the outcome under study is analysed outside of an instrumental variable framework. Searches included any papers published prior to 4 March 2022 in MEDLINE (PubMed), Embase (Ovid), Medline (Ovid) and MedRXiv. The search and full-text review were restricted to articles published in English. Outcome measures were any measure of health status or disease from a life stage after the exposure was measured. Study designs that do not use MR methods were not appraised. Treatment guidelines documents were excluded (Supplementary Material 2).

### 2. Data extraction and analysis

Within the final list of papers, we separated methodological manuscripts that presented or tested an approach to lifecourse MR from applied papers that only presented the results of a specific lifecourse analysis. For data collected from methodological manuscripts that presented or tested an approach to lifecourse MR we recorded: author, baseline year of data collection, aim, methodological approach, challenges in methodological application, simulation scenarios, sample size, and assumptions, and when an applied element was included in the manuscript, exposure, exposure age(s) in years, outcome and outcome age(s) in years were collected. We extracted the following from applied studies that presented the results of a specific lifecourse analysis: author, baseline year of data collection, aim, exposure, exposure age(s) in years, outcome and outcome age(s) in years. Title and abstract and then full-text screening was conducted in duplicate by two investigators (G.M.P and P.P.) and extraction in duplicate by two investigators (G.M.P and C.P.). Discrepancies were resolved by consensus. A narrative synthesis was performed. The evaluation of study quality by conducting a bias assessment was not considered relevant here, since we were not collating evidence to answer one applied question (20, 21).

## Results

Our search generated 317 published records. Two additional records were identified through conversations with experts in the field. After screening titles and abstracts, 135 manuscripts were assessed for eligibility. Of these, 97 articles were deemed eligible for inclusion in this systematic review (Figure 1). Thirteen studies presented or tested an approach to lifecourse MR (12-15, 22-30) and 84 presented the results of a specific lifecourse analysis without an emphasis on exploring or explaining a methodological approach (31–114). If a study fit the criteria for the former section, it was not included in the latter.

**Figure 1.**
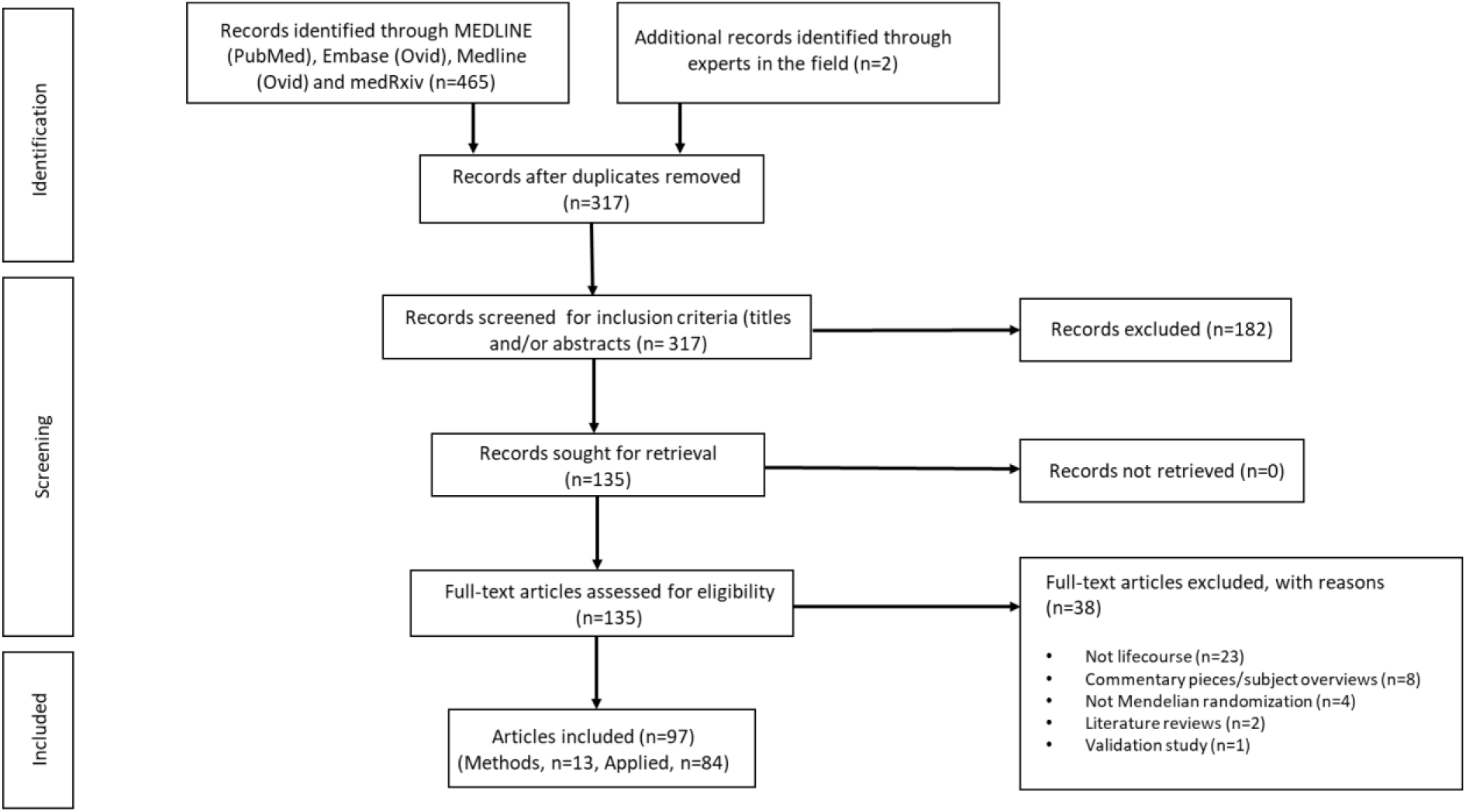
PRISMA flow chart illustrating selection of studies. PRISMA, Preferred Reporting Items for Systematic Reviews and Meta-Analyses

### Section 1: Studies presenting or testing an approach to lifecourse MR

Of the 13 studies presenting and/or testing approaches to lifecourse MR, four focused on the impact of time-varying exposures on the interpretations of “standard” MR techniques (12, 23, 26, 27). These additionally outline methods to assess and/or lessen potential bias. Five presented methods for analysing repeat measures of the same exposure. These comprised functional principal component (FPC) analysis through conditional expectation (PACE), multivariable MR (MVMR), G-estimation of structural nested cumulative failure models (SNCFTMs) and G-estimation of structural mean models (SMM) (13, 15, 22, 25, 28). Our definition of lifecourse studies, which includes the effects of repeated measures of the same time-varying exposure on a later outcome, connects lifecourse MR to G-estimation, which has been applied in several studies to adjust for time-varying confounding in traditional epidemiological settings (115, 116). Furthermore, four studies described novel methods that have been developed for intergenerational studies investigating a parental or grandparental exposure whilst the outcome of interest is assessed in offspring. These have used structural equation models (SEM) or the statistically equivalent linear mixed model (LMM) (14, 24, 29).

#### 1. Implications of time-varying exposures for the interpretation of “standard” MR

There are potential limitations regarding the use of “standard” MR techniques to interpret exposure-outcome relationships. D’Urso et al. (2020) highlight issues when using MR to assess the validity of hypotheses relating to the Developmental Origins of Health and Disease (DOHaD), such as the Barker hypothesis, which proposes that the origins of chronic diseases of adult life lie in foetal responses to the intrauterine environment (26). “Standard” MR methods do not take into account the relationship between maternal and offspring genotypes and, as a result, may produce inflated type 1 error rates. Standard errors may be too small in the presence of cryptic relatedness due to there being less genetic variation in the sample. A conditional analysis framework is recommended using an unweighted or weighted maternal allele score corrected for offspring genotypes (26).

Results from “standard” MR techniques are often interpreted as average lifetime effects of the exposure, i.e., the cumulative effect of the exposure level from conception and through the lifecourse. Labrecque et al. propose an alternative interpretation for exposures that vary over time. They suggest the effect should be interpreted using a counterfactual framework approach, shifting the entire exposure trajectory by one unit of time *k* (a timepoint of observation, where *k*=0 at conception) (23). Labrecque et al. argue that different effects would be estimated at different exposure time points if the relationship between the genetic variants and the exposure changes over time. Thus, a “standard” MR approach may produce biased results. They initially provided an empirical example to estimate the lifetime effect of body mass index (BMI) on systolic blood pressure using the rs9939609 variant. They then simulated a longitudinal relationship to estimate BMI as an exposure at age 30 and 50 years and concluded that when the genetic variable-exposure relationship was constant over time, estimates were unbiased with respect to the lifetime effect at both ages. In all other scenarios, however, they show the estimate differed, and this bias was sensitive to the strength of relationship between the genetic variant and exposure as well as the timing of measurement of both exposure window and outcome.

Previous studies have explored whether age modifies the relationship between the genetic variants and exposure (10), however, investigations are limited. Most studies that have addressed this have investigated body composition, BMI or other measures of body size. To assess how time-varying genetic effects may impact MR effect estimates, Labrecque et al. and others suggest looking at a statistical interaction between the genetic variant and age in relation to the exposure (13, 81, 82, 117–119). Following this, Labracque et al. propose plotting the relationship between the genetic instrument and the exposure stratified by age in samples with sufficient variation in age. They additionally show that patterns in age-varying genetic relationships may be exposure specific (27). This has been shown in applied studies (10, 13, 81, 82, 117–119).

Morris et al. clarify the causal estimates that are estimated by MR when applied to a single measure of a time-varying exposure with time-varying genetic effects (12). They consider a situation where there is one genetic instrument, a time-varying continuous exposure assessed on two occasions, and a single measure of an outcome. They also note the genetic instrument cannot affect the exposure measured at different occasions in isolation. Instead, they argue that the instrument underlies all possible exposure measurements across the lifecourse through a genetic liability, so a change in genotype changes both measures of the exposure. Simulations demonstrate that the Wald Ratio MR estimator recovers the correct causal effect in all scenarios assessed, even where time-varying genetic associations were present. Morris et al. claimed that MR estimates differ between measurements of time-varying exposures because MR is estimating the total effect of the exposure trajectory on the outcome rather than the effect of the exposure at a specific point in time. Further details of each of these approaches can be found in Supplementary Table 1.

#### 2. Methodological approaches to analysing repeat measures of the same exposure over the lifecourse in an MR framework

MR methods proposed to estimate the effects of repeat measures of the same exposure across the lifecourse have been developed in response to the concern that a single measurement of a time-varying exposure may not be adequate in capturing all time-varying information: a single measure of a time-varying exposure could underestimate the relationship between the exposure variable and the outcome variable, both due to the measurement error in the exposure and the failure to capture long-term change (120). Importantly, in this context, later stages of lifecourse exposures often depend on the earlier stages of the same exposure, whilst the reverse is not true.

Cao et al. developed two methods to combine functional data analysis (to describe the trajectory of the exposure) with MR, to test the causal effect of a time-varying exposure on a binary outcome (22). They use functional principal component (FPC) analysis through conditional expectation (PACE) to model the exposure trajectories, and then test whether a summary measure of the trajectory is related to the outcome using the two-stage residual inclusion (2SRI) approach. Their methods examine the evidence against the null hypothesis of no causal effect, but do not estimate the causal effect. The first method (PACE + 2SRI) assumes that the time-varying exposure variable has a cumulative effect on the risk of disease, and that the genetic effects on the exposure do not vary over time. The cumulative value of the exposure between two timepoints can be obtained by integration. The first stage obtains the residuals from regressing this cumulative exposure on the instrument (and any non-time- varying covariates). The second stage then relates these residuals to the outcome via a logistic regression model. For the second method (PACE+2SFRI), they allow a time-varying genetic effect on the exposure variable but assume that the effect of the exposure and the fitted residual on the outcome are constant over time. In this case, the first stage is a functional linear model for the time-varying exposure, and the second stage relates the outcome to the fitted residuals and to the detrended exposure (functional residual inclusion). The authors showed that this method outperformed “standard” MR analysis with a single measurement at one time point, with higher statistical power in simulation studies using the functional data analysis-based methods, even when the disease outcome was simulated to depend not on the cumulative exposure, but on the first three functional principal component scores from PACE.

Another method employed to assess repeat measures of the same exposure over the lifecourse is multivariable MR (MVMR) (13, 15). MVMR can be used to estimate the independent direct effects of several highly correlated exposures on an outcome, conditional on all the other exposures included in the model. It is useful in the context of mediation analysis (121), to estimate the effects of several repeated measures of the same exposure, or to isolate the effects of related phenotypes. Sanderson et al. explore the use of MVMR to estimate the direct effect of a single exposure at different time points in an individual’s lifetime on an outcome (Figure 2) (15). For multiple measurements to be included in a MVMR the genetic variants must have different effects on each exposure included in the model and these effects must not be a linear function of the others. The interpretation of the estimate is the effect of having a liability associated with a unit higher level of exposure at one occasion while keeping the liability for exposure at a separate occasion constant. Richardson et al. applied this approach to evaluate whether body size in early life has an independent effect on risk of disease in later life, or whether the effect seen is a result of body size in childhood being mediated by body size in adulthood (13). They use univariable MR to estimate total effects of early body size, and MVMR to estimate direct effects of early and adult body size. This approach suggests univariable analyses cannot identify critical or sensitive periods of exposure but can detect an effect of a difference in the cumulative lifetime exposure, which is a notion critiqued by Labrecque et al., highlighted earlier in this review (23, 27). If measures of the exposure at different time periods are available, and genetic instruments capable of reliably separating time-varying effects exist, it is possible to identify whether the exposure effects are stable over time or whether sensitive/critical periods exist in the lifecourse using MVMR. In theory the more timepoints we have should allow more granular inference into critical windows. However, whilst this method can narrow down or exclude periods, it cannot strictly identify important periods if the genetic effects on the periods included are correlated with genetic effects on excluded periods.

**Figure 2.**
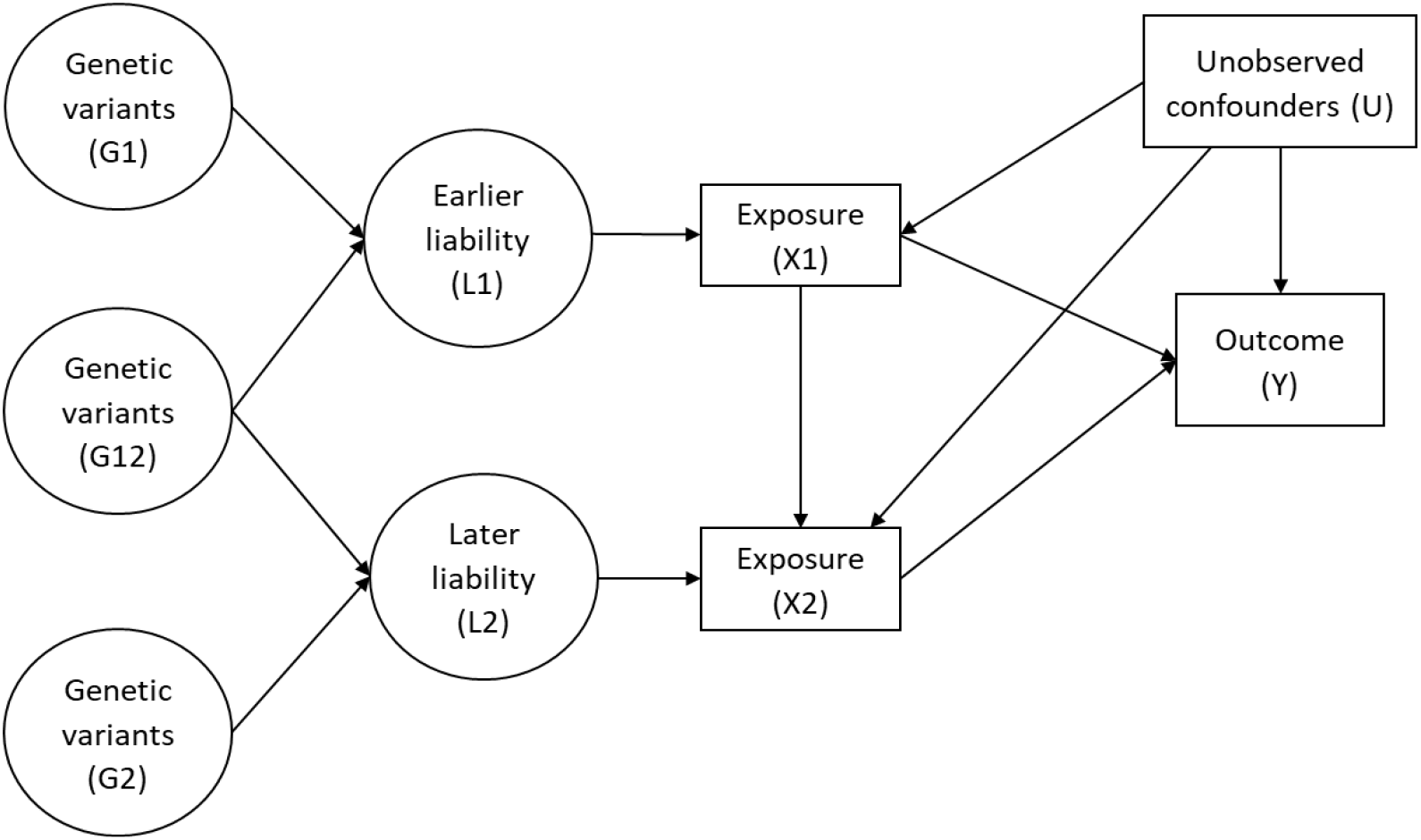
Latent exposure model with two periods of exposure (adapted from Sanderson et al.). *G*_1_ is a set of genetic variants associated with earlier liability (*L*_1_), *G*_2_ is a set of genetic variants associated with later liability (*L*_2_), *G*_12_ is a set of genetic variants associated with both *L*_1_ and *L*_2_.

The application of G-estimation of structural nested cumulative failure models (SNCFTMs) and G-estimation of structural mean models (SMM) was proposed by Shi et al. for the estimation of MR models with a time-varying exposure (Figure 3) (25, 28). Again, the interpretation of results from estimation for these models depends on the availability of data for the time-varying exposure. SNCFTMs can be used to estimate the causal effect of a time-varying treatment on a failure time outcome under the assumption that all time-varying confounders have been measured and that failure is rare under all possible treatment values (122). Shi et al. describe an adaptation of this use of SNCFTMs, incorporating IV-type assumptions (25). Whilst confirmation of the validity of the method was achieved via simulations, analyses indicated that MR with time-varying treatments and failure time outcomes using SNCFTMs require large sample sizes (n = 10,000; n = 25,000 or n = 50,000). In addition, authors note that this method should only be used with rare outcomes. In the application of g-estimation of SMMs to MR analyses, Shi et al. consider three types of causal effects that can be targeted when the exposure is time-varying: the effect of exposure at a single time point on the outcome (point effect), the effect of exposure during a period on the outcome (period effect), and the effect of exposure throughout the lifetime on the outcome (lifetime effect) (28). This approach highlighted two key challenges in estimating and interpreting period effects from MR analyses. The first is defining the period of interest. The second is the choice of time scale (e.g., time since conception or time since enrolment). In the context of additive causal effects for continuous outcomes, the authors note that g-estimation of SMMs and two-stage least squares (2SLS) MR yield similar estimates. SMMs can be naturally extended to many settings, including accommodating binary and failure-time outcomes and estimating effects on the multiplicative scale. SMMs are also semiparametric, and therefore avoid some of the parametric assumptions of 2SLS. Further details on these methodological approaches discussed along with their limitations are presented in Supplementary Table 1.

**Figure 3.**
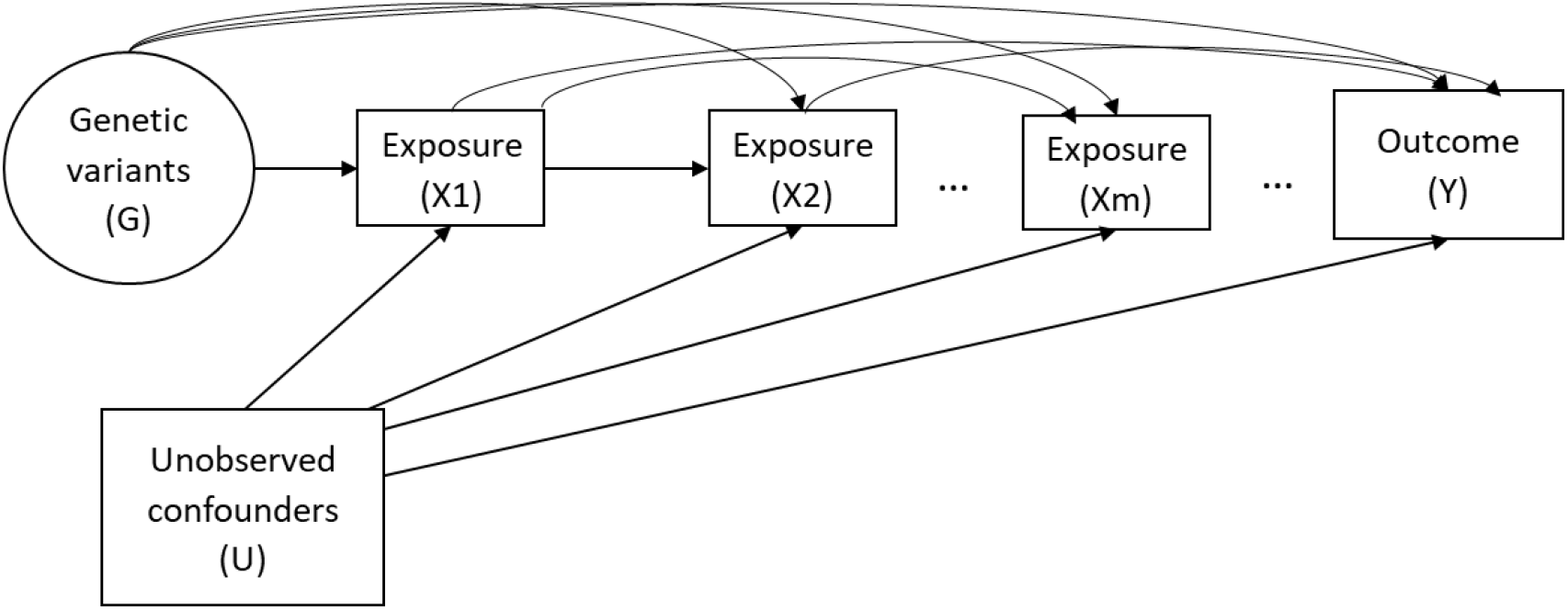
Causal diagram for instrumental variable analyses representing a scenario with a time-varying exposure (adapted from Shi et al. 2022).

#### 3. Novel methodological approaches to handling parental exposures in relation to offspring outcomes

Whilst “standard” MR approaches are largely sufficient for studies interested in estimating the causal effect of an early life exposure on a later life outcome, novel methods have been developed for intergenerational studies investigating a parental or grandparental exposure whilst the outcome of interest is assessed in offspring. All of the studies we identified in this section relate maternal genotypes to offspring outcomes and establish the causal effect of a maternal exposure, e.g., smoking during pregnancy, on offspring health. Yang et al. used a proxy gene-by-environment (G×E) MR approach to explore maternal effects on offspring phenotypes where maternal genetic information was unavailable (30). They validated this approach by replicating a known effect of maternal smoking heaviness on offspring birthweight using the rs16969968 variant in *CHRNA5*. They then applied it to explore effects of maternal smoking heaviness on offspring later life outcomes and on birthweight of participant’s children. Yang et al. demonstrated how G×E MR can be used to test transgenerational causal effects. Further studies included in this section emphasise the need to condition on offspring genotype to avoid including its effect on the outcome of interest. Earlier non-MR human genetic association studies have estimated maternal genetic effects on offspring phenotypes through conditional genetic association analysis of genotyped mother–offspring pairs (123). This separation of genetic effects into maternal and offspring components is important as maternal and offspring genotypes are correlated. Consequently, any association between maternal genotype and offspring outcome may be mediated by offspring genotype (Figure 4) (14, 29). Thus, as described above, naïve two-sample MR approaches in unrelated sets of individuals without accounting for the correlation between maternal and foetal genotype effects may result in erroneous conclusions regarding causality.

**Figure 4.**
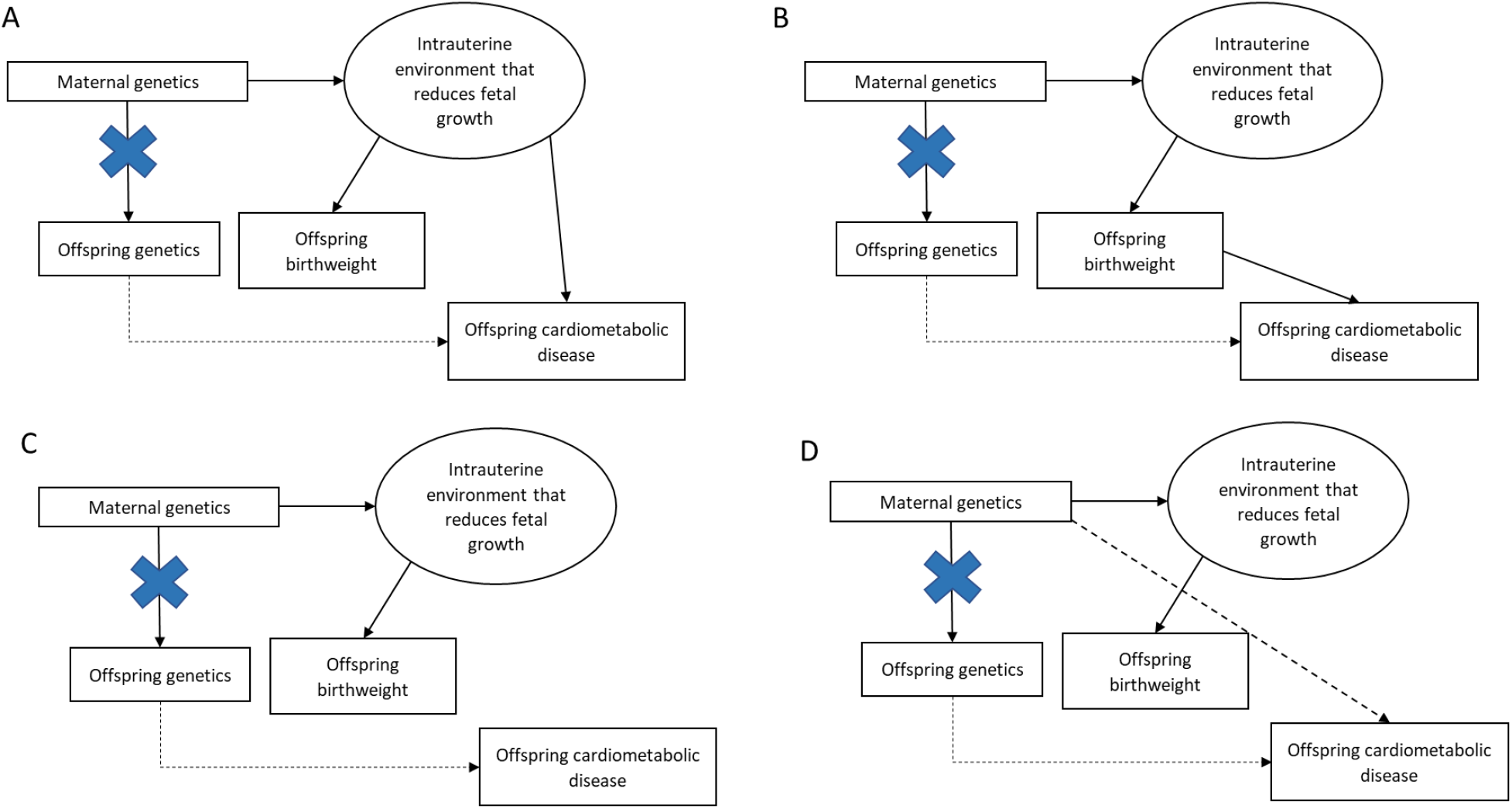
Four credible ways in which maternal single nucleotide polymorphisms (SNPs) can be related to offspring birthweight and offspring cardiometabolic risk factors. Blue crosses indicate the act of conditioning on maternal or offspring genotype, blocking the association between maternal and offspring variables. Dotted paths show paths in which the maternal genotype can be related to offspring phenotype that are not to do with intrauterine growth restriction (adapted from Moen et al.).

Two MR approaches, described by Warrington et al. and Evans et al. use structural equation modelling (SEM) to account for the correlation between maternal and foetal genotypes (14, 29). Evans et al. developed a statistical model that can be used to estimate the effect of maternal genotypes on offspring outcomes, conditional on offspring genotype using both individual-level and summary data. The authors demonstrate this approach using the following example: birthweight of the mothers, birthweight of the mother’s offspring, and the mother’s own genotype (SNP). The genotypes of the individual’s mother (their offspring’s grandmother) and the genotype of the individual’s offspring are considered latent unobserved variables. The total variance of the latent phenotype that relates to the genotype for the individual’s mother and offspring and for the observed SNP variable are estimated from the data. The causal path between the individual’s own genotype and both their mother and offspring’s latent genotype is set to 0.5, according to quantitative genetics theory. The estimated maternal and offspring effects of the SNP path coefficients, which refer to maternal and offspring genetic effects on birthweight, are also estimated. The resulting effects can be combined with SNP-exposure estimates for the maternal exposures that the investigator is interested in, in a two-sample MR framework.

Warrington et al. ran GWAS of own foetal genetic variants in relation to birthweight, and maternal genetic variants in relation to their offspring’s birthweight. They then partitioned the lead SNPs, representing independent association signals, into categories based on maternal and/or foetal genetic contributions to birth weight. To achieve this, they use SEM to account for the correlation between foetal and maternal genotypes to provide unbiased estimates of maternal and foetal genetic effects on birthweight. This method gives an indication as to which genetic associations are driven by the maternal and which by the foetal genomes. To extend the estimates of adjusted maternal and foetal effects genome wide, the authors developed a weighted linear model (WLM) which yields a good approximation of the SEM but is less computationally intensive. They used WLM-adjusted estimates in downstream analyses to identify foetal and maternal specific mechanisms that regulate birthweight and to investigate genetic links between birthweight and adult traits. Subsequently, the authors applied two-sample MR to estimate causal effects of intrauterine exposures on offspring birthweight. Authors selected SNPs associated with each exposure and regressed the WLM-adjusted maternal effects on birthweight for those SNPs against the effect estimates for the maternal exposure, weighting by the inverse of the variance of the maternal exposure effect estimates. Similarly, the authors used WLM-adjusted foetal effects to estimate the causal effect of the offspring’s genetic potential on their own birthweight and compare the results with the estimated maternal causal effects.

Moen et al. investigate whether a genetic risk score (GRS) of maternal SNPs associated with offspring birthweight is also associated with offspring cardiometabolic risk factors, after controlling for offspring GRS. They use a large dataset and explore father-offspring pairs to investigate whether there is evidence for a postnatal environmental effect (genetic nurture or dynastic effects) rather than an intrauterine environmental effect. In executing these analyses, the authors employ a LMM which accounts for the non-independence between siblings. They modelled offspring cardiometabolic risk factors as the outcome and included offspring birthweight, offspring birthweight squared, offspring age, offspring sex and measurement occasion as fixed effects. The non-independence between siblings and relatedness between parents and offspring was modelled using a genetic relatedness matrix in the random effects part of the model. This is described in detail (24). The authors performed primary analyses testing the relationship between maternal genetic risk scores (GRS) and each of the offspring risk factors, whilst conditioning on the offspring GRS. Further detail on applied results, assumptions and limitations for these methods are provided in Supplementary Table 1.

### Section 2: Applied MR studies presenting results of a lifecourse analysis

Of the 84 studies applying lifecourse MR methods, included in this review, 50% (42/84) estimated effects in just one generation, 45% (38/84) looked across two generations and 5% (4/84) estimating both. Of the one (and one and two) generational studies employed in this review, 59% (27/46) estimated the effect of exposures at birth, birth to/and childhood, birth to/and adolescence or birth to/and adulthood, 30% (14/46) at childhood, childhood to/and adolescence or childhood to/and adulthood, and 11% (5/46) at adolescence or adulthood. Within those focused on single generational effects, 50% (23/46) looked at birth weight, 46% (21/46) comprised body composition measures, including adiposity traits, BMI, body size, obesity, waist-to-hip ratio, and body fat percent. Single generation studies additionally included estimating the genetically predicted effects of age at menarche, pubertal age (timing), first sexual intercourse, sleep duration, offspring fasting glucose and type 2 diabetes, genetic liability to juvenile idiopathic arthritis, disordered eating pattern, alcohol consumption and DNA methylation at the HLA locus. Amongst the studies that estimated effects across two generations, 21% (9/42) examined body composition as exposure measures. These included maternal and paternal BMI as well as maternal adiposity, central obesity, and height. Other exposures examined in a two-generational setting are included in Supplementary Table 2. All of the two-generational studies estimated effects of maternal exposures, with one study also examining paternal exposures (124). Outcomes addressed in the studies incorporated in this review are varied and can be found in Supplementary Table 2.

## Discussion

In this systematic literature review, we extracted and summarised findings from studies presenting and/or testing approaches to lifecourse MR as well as those presenting results of a specific lifecourse analysis. Among the former, we focused on papers addressing time-varying or lifecourse processes through interpretations of results from “standard” MR techniques. “Standard” MR techniques have focused on estimating lifetime effects of an exposure, i.e., the cumulative effect of the exposure level from conception and through the lifecourse. Labrecque et al. propose that MR estimates of the same exposure assessed at different ages vary in the presence of time-varying genotype-exposure associations, and this represents bias in estimates of a lifetime causal effect. In response, Morris et al. proposed that “standard” MR is not estimating the causal effect of an exposure as it manifests at a given timepoint, but the causal effect of the underlying exposure liability. Thus, a hypothetical change in genotype would affect all manifestations of the exposure. In addition, papers comprising methodological approaches for intergenerational effects or pregnancy/birth exposures emphasised the importance of a statistical model that can estimate the effect of maternal genotypes on offspring outcomes, conditional on offspring genotype. On a related note, carrying out MR of own birthweight using only genetic variants of the individual is likely to result in inaccuracies. This is because foetal growth and subsequently birthweight may be influenced by both foetal and correlated maternal genotypes (57).

We summarised papers employing a methodological approach for repeat measures of the same exposure over the lifecourse. The methods described here enhance capability for causal inference of lifecourse effects, however, there are clear limitations. One method comprised the FPC analysis through PACE. Authors generated this for hypothesis testing, not for causal effect size estimation, and thus this may not provide consistent estimates. Another technique utilised was MVMR, which can separate influences across the lifecourse under some but not all causal scenarios. Moreover, estimates used in the studies presented are based solely on body size and BMI data from the UK Biobank (125, 126). These findings need to be evaluated in future cohorts when sample sizes make this possible. This is particularly important as it has been shown that UK Biobank participants are highly selected, which can be problematic for instrumental variables analyses (125, 126). In addition, a G-estimation of SNCFTMs was explored. If the rare failure assumption does not hold, however, estimates from this approach may be invalid. Informative MR analyses will additionally require sample sizes much larger than those presented in this paper. A G-estimation of SMM was also described. Due to wide variations in age at first visit and short duration of follow-up in the data used, authors were limited to using time since enrolment in the study as the time scale, which implies the added assumption that the period effect is homogeneous across age. The plausibility of this assumption is not only specific to the exposure–outcome relationship of interest, but also depends on the variability in age.

Among the studies presenting results of specific lifecourse analyses, data availability limitations were apparent. Studies focusing on one generational research are largely confined to the exploration of questions regarding body composition, since these have the strongest instrumental variables. In addition, these data are often more commonly available on a large scale in most longitudinal cohorts. This emphasizes the need for pooling data across studies to maximise power, highlighting the value of a Lifecourse MR consortium, which will enable the testing of key epidemiological hypotheses that have been advanced regarding critical period and cumulative effects on disease risk. For some phenotypes, however, lifecourse MR may not be able to usefully contribute. This could either be due to the lack of identified genetic variants allowing meaningful separation of measures at different life stages or because these simply do not exist. If the effects IV-exposure effects are relatively constant, “standard” MR may therefore be sufficient. Awareness of this may change over time as more data becomes available. The collection of these data is likely to be useful to improve MR overall. For example, stratifying analyses by age could be of value for testing other MR assumptions. An instrument that has very little effect on the earlier life exposure whilst influencing a later-life exposure and associating with an early-life outcome may be indicative of violations of horizontal pleiotropy, correlated pleiotropy, as well as the gene-environment equivalence (‘consistency’) assumption. In addition, lifecourse data may be used for evidence of substantial *in utero* effects of variants on processes suggesting developmental trajectories. Future work is required to develop guidance on how best to implement MR methods with the data that may be available, within a lifecourse epidemiology framework.

## Conclusions

There is a growing body of research focused on the development of lifecourse MR techniques and methods which are increasingly being applied to address lifecourse research questions. The possibility that genetic effects have different levels of importance in the development of an exposure at different time points should be more commonly considered for application when conducting MR investigations. Clear limitations persist, however, specifically regarding data constraints.

## Data Availability

All data relevant to the study are included in the article or uploaded as online supplemental information.

## Statements & Declarations

### Ethics approval

This study is a systematic review of available literature and did not involve direct access to participants of the primary research studies included. Research ethics approval was therefore not required.

### Consent to participate

Not applicable.

### Authors’ contributions

G.M.P: Conceptualisation, Methodology, Software, Formal analysis, Investigation, Data curation, Writing – original draft, Writing – review and editing, Visualisation. E.S: Writing – review and editing. P.P: Validation, Writing - review and editing. A.F: Writing – review and editing. T.T.M: Writing – review and editing. C.P: Validation. T.M.F: Writing – review and editing. J.H: Writing – review and editing. T.G.R: Writing – review and editing. R.R: Conceptualisation. J.T: Writing – review and editing. G.D.S: Conceptualisation, Writing – review and editing, Supervision, Funding acquisition. L.D.H: Conceptualisation, Writing – review and editing, Supervision. K.T: Conceptualisation, Writing – review and editing, Supervision.

