## Supplementary Material 1 for "A systematic literature review of methodological approaches, challenges, and opportunities in the application of Mendelian randomisation to lifecourse epidemiology"

**Supplementary Material 1.** Search strategy used to identify lifecourse Mendelian randomisation studies

MEDLINE (PubMed)

("lifecourse"[Title/Abstract] OR "life course"[Title/Abstract] OR "time-varying"[Title/Abstract] OR "childhood adult"[Title/Abstract] OR (("childhood"[All Fields] OR "childhoods"[All Fields]) AND "later life"[Title/Abstract]) OR ("early"[All Fields] AND "later life"[Title/Abstract]) OR ("early"[All Fields] AND "life adult"[Title/Abstract]) OR (("birth s"[All Fields] OR "birthed"[All Fields] OR "birthing"[All Fields] OR "parturition"[MeSH Terms] OR "parturition"[All Fields] OR "birth"[All Fields] OR "births"[All Fields]) AND ("adult"[MeSH Terms] OR "adult"[All Fields] OR "adults"[All Fields] OR "adult s"[All Fields])) OR (("birth s"[All Fields] OR "birthed"[All Fields] OR "birthing"[All Fields] OR "parturition"[MeSH Terms] OR "parturition"[All Fields] OR "birth"[All Fields] OR "births"[All Fields]) AND "later"[All Fields] AND ("life"[MeSH Terms] OR "life"[All Fields])) OR (("gestate"[All Fields] OR "gestated"[All Fields] OR "gestates"[All Fields] OR "gestating"[All Fields] OR "gestational"[All Fields] OR "gestations"[All Fields] OR "pregnancy"[MeSH Terms] OR "pregnancy"[All Fields] OR "gestation"[All Fields]) AND ("adult"[MeSH Terms] OR "adult"[All Fields] OR "adults"[All Fields] OR "adult s"[All Fields])) OR (("gestate"[All Fields] OR "gestated"[All Fields] OR "gestates"[All Fields] OR "gestating"[All Fields] OR "gestational"[All Fields] OR "gestations"[All Fields] OR "pregnancy"[MeSH Terms] OR "pregnancy"[All Fields] OR "gestation"[All Fields]) AND "later"[All Fields] AND ("life"[MeSH Terms] OR "life"[All Fields])) OR (("uterus"[MeSH Terms] OR "uterus"[All Fields] OR "utero"[All Fields]) AND ("adult"[MeSH Terms] OR "adult"[All Fields] OR "adults"[All Fields] OR "adult s"[All Fields])) OR (("uterus"[MeSH Terms] OR "uterus"[All Fields] OR "utero"[All Fields]) AND "later"[All Fields] AND ("life"[MeSH Terms] OR "life"[All Fields])) OR (("intrauterin"[All Fields] OR "intrauterine"[All Fields]) AND ("adult"[MeSH Terms] OR "adult"[All Fields] OR "adults"[All Fields] OR "adult s"[All Fields])) OR (("intrauterin"[All Fields] OR "intrauterine"[All Fields]) AND "later"[All Fields] AND ("life"[MeSH Terms] OR "life"[All Fields])) OR (("fetale"[All Fields] OR "fetally"[All Fields] OR "fetals"[All Fields] OR "fetus"[MeSH Terms] OR "fetus"[All Fields] OR "fetal"[All Fields] OR "foetal"[All Fields]) AND ("adult"[MeSH Terms] OR "adult"[All Fields] OR "adults"[All Fields] OR "adult s"[All Fields])) OR (("fetale"[All Fields] OR "fetally"[All Fields] OR "fetals"[All Fields] OR "fetus"[MeSH Terms] OR "fetus"[All Fields] OR "fetal"[All Fields] OR "foetal"[All Fields]) AND "later"[All Fields] AND ("life"[MeSH Terms] OR "life"[All Fields])) OR (("fetale"[All Fields] OR "fetally"[All Fields] OR "fetals"[All Fields] OR "fetus"[MeSH Terms] OR "fetus"[All Fields] OR "fetal"[All Fields] OR "foetal"[All Fields]) AND ("adult"[MeSH Terms] OR "adult"[All Fields] OR "adults"[All Fields] OR "adult s"[All Fields])) OR (("fetale"[All Fields] OR "fetally"[All Fields] OR "fetals"[All Fields] OR "fetus"[MeSH Terms] OR "fetus"[All Fields] OR "fetal"[All Fields] OR "foetal"[All Fields]) AND "later"[All Fields] AND ("life"[MeSH Terms] OR "life"[All Fields])) OR (("offspring"[All Fields] OR "offspring s"[All Fields] OR "offsprings"[All Fields]) AND ("pregnancy"[MeSH Terms] OR "pregnancy"[All Fields] OR "pregnancies"[All Fields] OR "pregnancy s"[All Fields])) OR (("offspring"[All Fields] OR "offspring s"[All Fields] OR "offsprings"[All Fields]) AND ("maternally"[All Fields] OR "maternities"[All Fields] OR "maternity"[All Fields] OR "mothers"[MeSH Terms] OR "mothers"[All Fields] OR "maternal"[All Fields])) OR ("intergeneration"[All Fields] OR "intergenerational"[All Fields]) OR "two-generational"[All Fields]) AND ("mendelian randomization"[Title/Abstract] OR "mendelian randomisation"[Title/Abstract] )

- 184 results

MedRXiv

("lifecourse" OR "life course" OR "time-varying”) AND ("mendelian randomization” OR "mendelian randomisation")

- 51 results

Medline (Ovid)

((lifecourse or life course or time-varying or childhood adult or childhood later life or early later life or early life adult or birth adult or birth later life or fetal adult or fetal later life or foetal adult or intrauterine later life or intrauterine adult) and (mendelian randomization or mendelian randomisation))

- 103 results

Embase (Ovid)

((lifecourse or life course or time-varying or childhood adult or childhood later life or early later life or early life adult or birth adult or birth later life or fetal adult or fetal later life or foetal adult or intrauterine later life or intrauterine adult) and (mendelian randomization or mendelian randomisation))

- 127 results

Total: 465 results

Duplicates: 150

Remaining 315 + 2 additional records identified through other sources
