## Supplementary Material 2 for "A systematic literature review of methodological approaches, challenges, and opportunities in the application of Mendelian randomisation to lifecourse epidemiology"

**Supplementary Material 2.** Inclusion and exclusion criteria

| **Inclusion Criteria** |  |
| --- | --- |
| Types of studies | Publication date: ≤ 4 March 2022 |
|  | Studies from any geographical location. |
|  | English publication language |
|  | Studies using Mendelian randomisation methods |
|  | Lifecourse epidemiology studies, defined as studies investigating biological, behavioral, and psychosocial processes that link health and disease risk later in life to exposures that take place in a preceding life stage (pre-gestation, gestation, early life, childhood, adolescence) to the outcome, or the effects of repeated measures of a time-varying exposure on a later outcome. |
| Types of participants | All acceptable. |
| Types of exposure measures | Physical or social exposures measured on one or more than one occasion during gestation, childhood, adolescence, earlier or adult life or across generations as long as at least one measure pertains to a life stage before outcome is measured. |
| Types of outcome measures | Any health status or disease risk later in life, defined as a measure taken from a life stage after the exposure was measured. |

| **Exclusion Criteria** |  |
| --- | --- |
| Types of studies | Exclusively observational study designs that do not use Mendelian randomisation methods |
|  | Treatment guidelines documents, other reviews |
